# Glycemic response trajectories on metformin monotherapy in real-world diabetes care

**DOI:** 10.64898/2026.05.24.26353996

**Authors:** Sridharan Raghavan, Wenhui Liu, Max R. Ho, Theodore Warsavage, Debashis Ghosh, Liron Caplan, Jane E. B. Reusch

**Affiliations:** US Department of Veterans Affairs, Eastern Colorado Health Care System, Aurora, CO; Department of Medicine, University of Colorado Anschutz Medical Campus, Aurora, CO; Department of Biostatistics and Informatics, Colorado School of Public Health, Aurora, CO

## Abstract

**Objectives:** Diabetes affects over 500 million people globally and glycemia is inadequately managed. Metformin is the most frequently prescribed initial treatment for type 2 diabetes globally, yet glycemic response trajectories to metformin in routine real-world care and predictors of treatment response have not been well described. We aimed to identify glycemic response trajectories in adults prescribed metformin monotherapy as initial type 2 diabetes treatment and predictors of poor glycemic response to metformin.

**Design:** Observational cohort study using latent class mixed models to identify hemoglobin A1c (HbA1c) trajectory classes, followed by random forests machine learning to predict trajectory class membership.

**Setting:** US Veterans Affairs Healthcare System

**Participants:** Adults treated with metformin alone for >30 days after diabetes diagnosis with a minimum of two HbA1c measurements from 90 days prior to two years after the first metformin prescription (N=140,413).

**Exposures:** Demographic, laboratory, vital sign, and comorbidity data were included as predictors of metformin response trajectory

**Main Outcomes and Measures:** We included all HbA1c measurements (487,604 total) for two years after metformin initiation to define metformin glycemic response trajectories.

**Results:** We identified three HbA1c trajectories: stably low (89.7% of sample, mean HbA1c decrease from 7.2% to 6.6%), brisk response (7.1% of sample, mean HbA1c decrease from 11.4% to 7.0%), and non-response (3.1% of sample, mean HbA1c increase from 8.9% to 10.8%). Of those in the stably low and brisk response classes at 2 years, 91% maintained HbA1c at approximately 7% on metformin alone for 5 years after drug initiation. Prediction models could accurately predict brisk response (91% accuracy) but not metformin non-response (59% accuracy).

**Conclusions:** Most individuals treated initially with metformin monotherapy have a beneficial and durable glycemic response. Predicting individuals who will not respond to metformin may be challenging but is evident within six months with recommended glycemic surveillance. The findings support current guidelines for HbA1c surveillance when initiating diabetes treatment.

**Summary Box:** 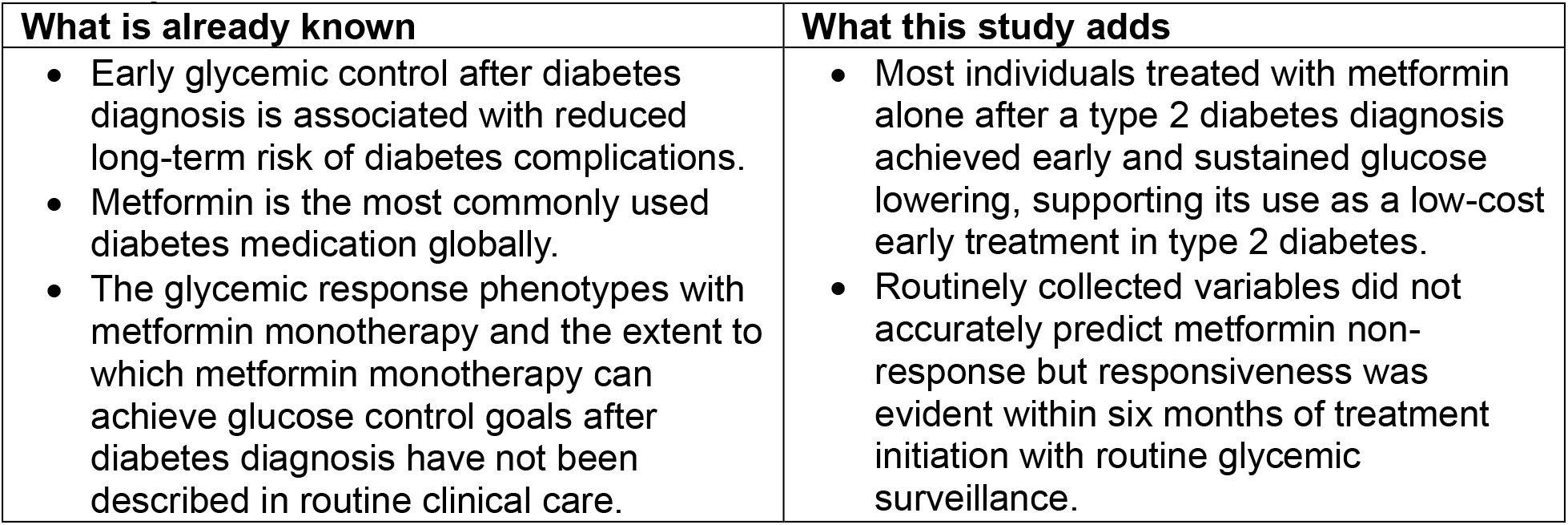

## Introduction

Type 2 diabetes affects over 10% of the US population^1^. Measures of diabetes care suggest low levels of achievement of guideline recommended treatment goals, including glycemic control, blood pressure, and cholesterol reduction^2,3^. While recent attention has focused on new classes of type 2 diabetes medications with cardiovascular and kidney benefits^4^, early glycemic control after diabetes diagnosis is another proven strategy for reducing risk of diabetes complications^5^. Real-world data on initial diabetes treatment response is scarce.

Metformin is a safe, low-cost treatment for type 2 diabetes and is the most frequently prescribed initial medication after a diagnosis of type 2 diabetes in the US and worldwide^6,7^. However, few clinical trials have studied initial medication choice in type 2 diabetes, and evidence to guide initial diabetes treatment in real-world diabetes patient populations is limited. The UK Diabetes Prospective Study (UKPDS) and A Diabetes Outcome Progression Trial (ADOPT) randomized individuals with type 2 diabetes to different first-line diabetes treatment strategies and evaluated glycemic response^5,8-10^. The trials had narrow inclusion/exclusion criteria and were designed to examine glycemic response as a binary outcome – achievement of a target for glycemic control (UKPDS) or treatment failure based on glucose exceeding a specified threshold (ADOPT). The translation of these clinical trials to initial treatment selection in real-world populations with newly diagnosed diabetes, particularly whether metformin may be an appropriate or inappropriate initial medication, remains challenging.

We aimed to describe the natural history of glycemia in individuals initially treated with metformin monotherapy using longitudinal trajectory models. Because metformin predominates as the initial treatment for type 2 diabetes worldwide, a secondary aim of this study was to develop a model to predict glycemic response to metformin, particularly to identify individuals likely to have a poor glycemic response to metformin who may benefit from alternative initial treatment. Using real-world data on early glycemic control on metformin, we seek to provide insights on strategies to optimize long-term outcomes in individuals with type 2 diabetes through prompt and effective glucose lowering with low-cost treatment.

## Methods

### Study participants

We have previously described the criteria for identifying a cohort of individuals with newly diagnosed diabetes in the course of routine primary care in the VA healthcare system based on a validated algorithm for the VA^11,12^. Briefly, diabetes status was defined as ≥2 uses of ICD-9-CM diagnosis codes 250.xx or ≥1 use of 250.xx associated with a primary care provider (PCP) visit in conjunction with an outpatient diabetes medication prescription. To identify newly diagnosed individuals receiving initial diabetes treatment, participants had to meet two criteria in the two years prior to the above diabetes diagnosis definition: 1. A minimum of two clinical encounters in the VA, one of which had to be a PCP encounter; 2. At least one prescription medication filled in the VA. These conditions result in a cohort with evidence of active VA care without recognized diabetes or diabetes treatment for a look back period of two years prior to meeting the diabetes diagnosis criteria. We included individuals diagnosed with diabetes based on the above criteria and receiving initial monotherapy between 2005 and 2016. Study entry was limited to 2016 to permit analysis of up to five years of follow-up data (through 2021).

Initial diabetes treatment was based on prescription medication fills occurring on or after the date of the first occurrence of a diabetes ICD-9-CM diagnosis code. Individuals prescribed metformin prior to the first occurrence of a diabetes diagnosis code were excluded as were individuals with prescriptions for metformin or other diabetes medications in non-VA care during the first two years after diabetes diagnosis. Initial metformin monotherapy was defined as a filled prescription for metformin without a filled prescription for any other diabetes medication or metformin combination medication within the subsequent 4 weeks. The date of the initial metformin prescription fill was considered the date of treatment initiation and served as the baseline date for an individual study participant. The Colorado Multiple Institutional Review Board and the Research & Development Committee of the VA Eastern Colorado Healthcare System provided human subjects oversight and approval.

### Glycemic trajectory modeling

Hemoglobin A1c (HbA1c) trajectories were based on all HbA1c measurements occurring from 90 days before the baseline date through two- or five-years after baseline. To be included, individuals had to have at least two HbA1c measurements in the specified time window. Individuals were censored if they were prescribed a second diabetes medication, were admitted to a long-term care facility, had 90-day metformin adherence of less than 80%, or died. Metformin adherence was estimated as the proportion of days covered based on VA pharmacy fill data for each 90-day period after baseline^13^. We used latent class mixed models (R package *lcmm*) to identify different HbA1c trajectory classes after initial metformin treatment until censoring or the end of the follow-up period. The primary analysis included two years of follow-up, and secondary analysis included five years of follow-up. We found that a third-degree polynomial of time, with a random effect for time, best fit the data. We fit models with one to six latent trajectory classes and evaluated model fit using the Bayes Information Criterion (BIC) and trajectory class size based on posterior probability of class membership. Finally, as a sensitivity analysis, we repeated the two- and five-year trajectory models without censoring for metformin non-adherence.

### Prediction models for metformin response

For ease of analysis and interpretability, we focused on the three-class glycemic trajectory model for identifying predictors of metformin response as it included trajectory classes exhibiting HbA1c lowering and increasing on metformin. Based on the three-class model for two-year HbA1c trajectory, we compared baseline patient characteristics across different trajectory classes using chi-square tests to compare categorical data and Mann-Whitney Wilcoxon nonparametric tests for continuous data.

To develop a prediction model for HbA1c trajectory class, we evaluated three sets of predictors: first with just five variables (baseline age, HbA1c, body mass index [BMI], triglycerides [TG], and high-density lipoprotein cholesterol [HDL]), second with 10 variables (addition of baseline alanine aminotransferase [ALT], aspartate aminotransferase [AST], and presence/absence of three comorbidities -heart failure [HF], coronary artery disease [CAD], and liver disease), and finally with twenty-three additional variables for baseline comorbidities or medication use (listed in **Supplemental Table 1**). The data were divided randomly into two-thirds for training and one-third for testing. We developed prediction models using random forests, a machine learning algorithm that uses a series of decision trees that accommodate interactions to prioritize variables that predict the outcome, in this case, metformin trajectory class ^14^. Internal validation for all models was conducted using 10-fold cross validation repeated three times. Prediction model accuracy was assessed using a confusion matrix to estimate accuracy and the kappa statistic for classification into each trajectory class^15^. The kappa statistic is a more useful measure of prediction accuracy in this study application as it accounts for unbalanced proportions of individuals in each of the three trajectory classes. As a sensitivity analysis, we repeated the prediction model development using neural networks and support vector machines (SVM), two additional machine learning algorithms widely used for classification^16,17^.

## Results

A total of 140,413 individuals with newly diagnosed diabetes treated with metformin monotherapy were included. They were predominantly male (96%) with a mean age of 62 years, mean HbA1c of 7.5%, and mean BMI of 33.7 kg/m^2^ at baseline (**Table 1**). Common comorbidities at baseline include a history of cancer, coronary artery disease (CAD), arthritis, depression or anxiety, hyperlipidemia, and hypertension (**Table 1**).

When modeling two-year glycemic response trajectories on metformin, the BIC improved as the number of trajectory classes increased from two to six (**Supplemental Table 2**). Three distinct trajectory classes emerged in the three-class model (**Figure 1A**): individuals with a low HbA1c at treatment initiation who maintained a low HbA1c over follow-up (Class 1, 89.7%), those with a high HbA1c at treatment initiation who had a brisk HbA1c decline within six months (Class 2, 7.1%), and those with an intermediate HbA1c at treatment initiation that increased over the follow-up period (Class 3, 3.1%). Models with three to six trajectory classes consistently included a class with >85% of the study sample resembling Class 1, and a class with >5% of the study sample resembling Class 2. While increasing the number of classes in the model improved the BIC, implying better model fit to the data, the more complex models resulted in additional classes with only small proportions of the study sample (**Supplemental Table 3; Supplemental Figure 1**). Accordingly, we focused the remainder of the analyses on the three-class model that included individuals who did and did not have a favorable initial glycemic response to metformin treatment.

**Table 1.**
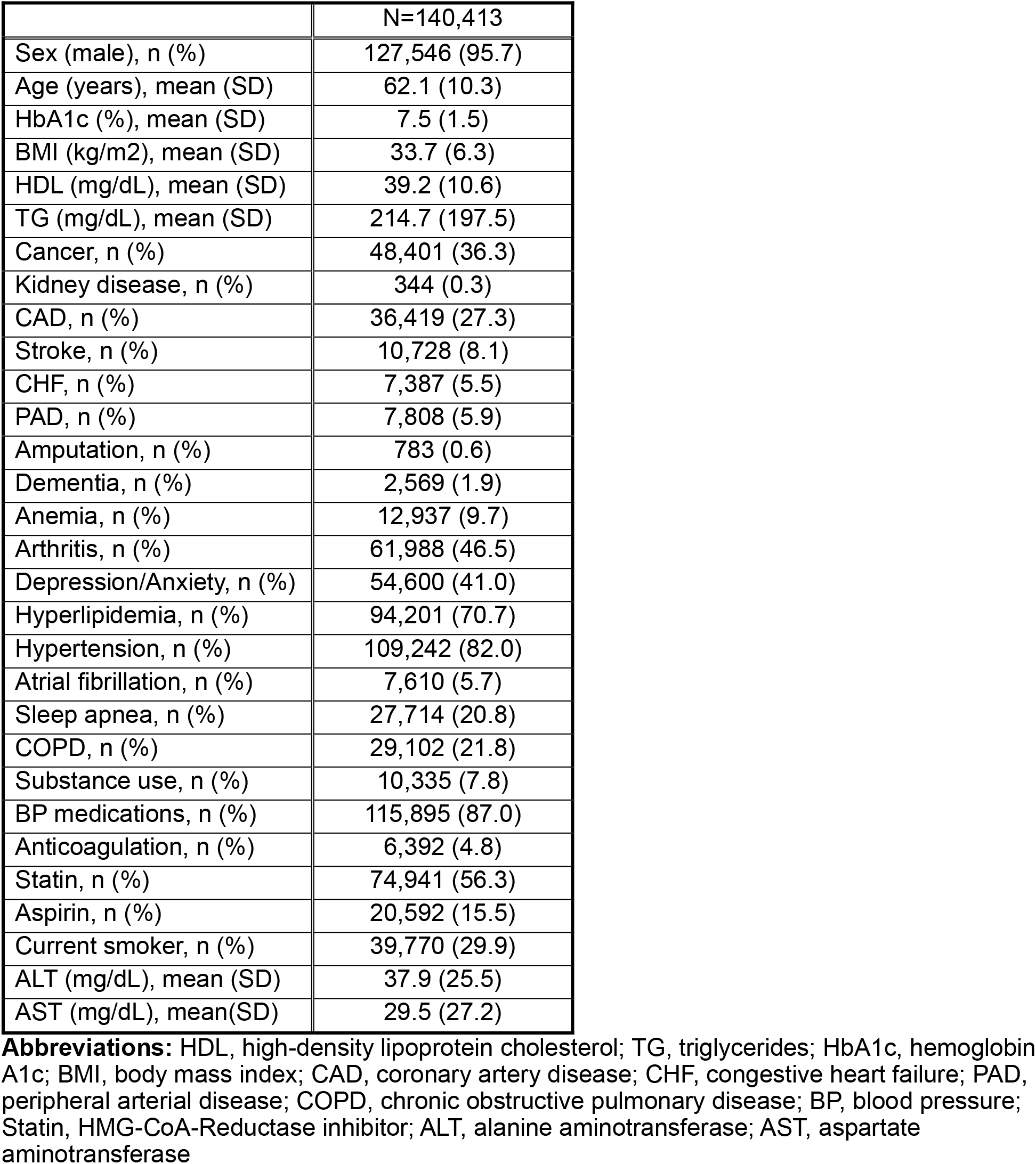
Baseline characteristics of study participants.

**Figure 1.**
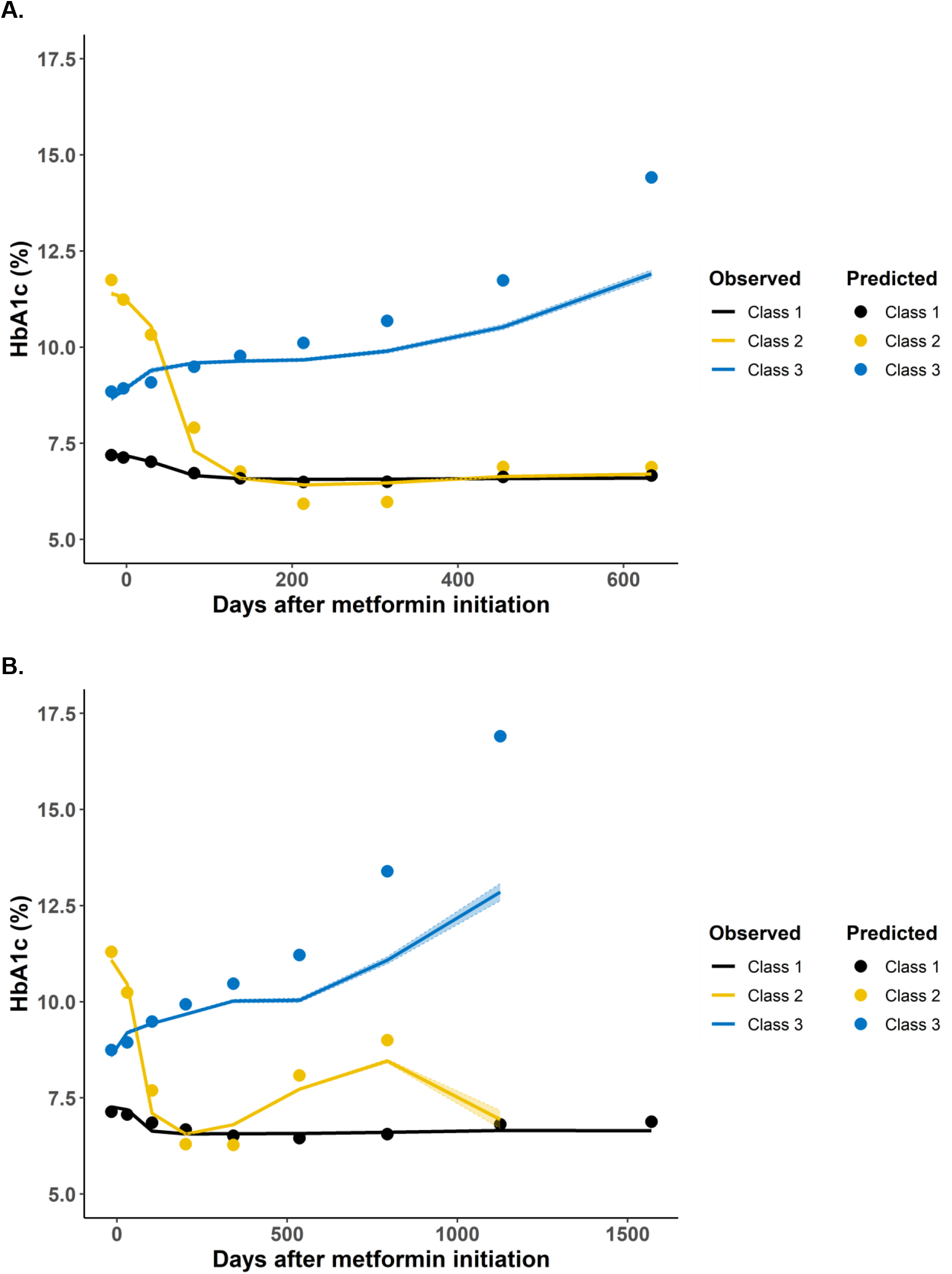
**(A)** Two-year hemoglobin A1c (HbA1c) trajectories classified with a three-trajectory model. **(B)** Five-year hemoglobin A1c (HbA1c) trajectories classified with a three-trajectory model. Observed HbA1c trends and confidence intervals are represented by lines and shading; predicted HbA1c based on trajectory model are represented by points. Individuals were censored for initiation of a second diabetes medication, death, admission to a long-term care facility, or <80% adherence to metformin in a 90-day period. All individuals classified into Classes 2 and 3 were censored prior to the end of the five-year follow-up period in (B).

When follow-up time was extended to five-years in a three-class model, the majority of individuals were classified into a trajectory with low HbA1c at treatment initiation and remained well-controlled over the follow-up period (Class 1, 91.7% of the sample) (**Figure 1B**). A second class with high baseline HbA1c with a brisk positive initial glycemic response again fit the data well (Class 2, 5.2% of the sample); this second class exhibited glycemic deterioration after two to three years and was mostly censored by the fourth year of follow-up. Finally, a third class with intermediate baseline HbA1c that gradually increased while on metformin monotherapy was again seen when five years of follow-up data were used to fit the trajectory class models (Class 3, 3.1% of the sample).

Censoring over the first two years of follow-up was common in all three classes (**Supplemental Table 4**). Metformin non-adherence was the most common cause of censoring in Classes 1 and 2, and initiation of a second diabetes medication was the most common cause of censoring in Class 3 (**Supplemental Table 4**). Death and long-term care facility admission were rare causes of censoring across all classes. When individuals were not censored for metformin non-adherence, 84% of the participants remained in the analysis through two years of follow-up. The results of the three-class model were stable in a sensitivity analysis in which metformin adherence was not required to remain in the analysis: 84.2% of individuals were classified into Class 1, 8.3% into Class 2, and 7.6% into Class 3 (**Supplemental Figure 2**).

Baseline characteristics differed between individuals assigned to each trajectory class (**Table 2**). Class 1 individuals were older and had higher prevalence of most comorbidities examined. Class 3 individuals (non-responders) had the youngest age on average, the highest BMI, and the highest prevalence of depression/anxiety and active smoking (**Table 2**). Random forests prioritized HbA1c and age as the first and second most important variables for predicting metformin trajectory class membership (**Supplemental Table 5**). Prediction model accuracy for assignment to one of three glycemic response trajectories was high (accuracy 0.94 and kappa 0.67; **Table 3**), suggesting substantial agreement in trajectory class prediction and observed assignment in the held-out testing/validation sample. However, prediction accuracy can be inflated when the proportions of individuals in each class are unbalanced, as in this study, where nearly 90% of participants were assigned to Class 1. Examining class-specific performance revealed that accuracy of predicting glycemic non-response to metformin was poor (<0.6) irrespective of the number of predictors included in model development or the machine learning approach used (**Table 3, Supplemental Tables 6 and 7**). Overall, the model performed well in accurately classifying into the majority class but performed poorly in identifying the relatively small subgroup of metformin non-responders.

**Table 2.** Baseline characteristics of study participants stratified by trajectory classification.

|  | **Class 1**<br>N=125,981 | **Class 2**<br>N=10,021 | **Class 3**<br>N=4,411 | **p** |
| --- | --- | --- | --- | --- |
| Sex (male) , n (%) | 114,615 (95.6) | 9,058 (96.8) | 3,873 (96.5) | <0.001 |
| Age (years), mean (SD) | 62.6 (10.1) | 58.2 (10.4) | 56.1 (10.5) | <0.001 |
| HbA1c (%), mean (SD) | 7.2 (0.88) | 11.4 (1.4) | 8.9 (1.7) | <0.001 |
| BMI (kg/m<sup>2</sup>), mean (SD) | 33.7 (6.3) | 33.0 (5.9) | 35.6 (6.9) | <0.001 |
| HDL (mg/dL), mean (SD) | 39.4 (10.6) | 37.0 (10.2) | 37.6 (10.2) | <0.001 |
| TG (mg/dL), mean (SD) | 204.0 (165.2) | 316.4 (385.5) | 295.6 (317.7) | <0.001 |
| Cancer, n (%) | 44,370 (37.0) | 2,868 (30.6) | 1,163 (29.0) | <0.001 |
| Kidney disease, n (%) | 317 (0.3) | 16 (0.2) | 11 (0.3) | 0.224 |
| CAD, n (%) | 33,811 (28.2) | 1,756 (18.8) | 852 (21.2) | <0.001 |
| Stroke, n (%) | 9,945 (8.3) | 545 (5.8) | 238 (5.9) | <0.001 |
| CHF, n (%) | 6,800 (5.7) | 360 (3.8) | 227 (5.7) | <0.001 |
| PAD, n (%) | 7,219 (6.0) | 389 (4.2) | 200 (5.0) | <0.001 |
| Amputation, n (%) | 702 (0.6) | 50 (0.5) | 31 (0.8) | 0.245 |
| Dementia, n (%) | 2,362 (2.0) | 135 (1.4) | 72 (1.8) | 0.001 |
| Anemia, n (%) | 11,925 (10.0) | 671 (7.2) | 341 (8.5) | <0.001 |
| Arthritis, n (%) | 56,495 (47.1) | 3,778 (40.4) | 1,715 (42.7) | <0.001 |
| Depression/Anxiety, n (%) | 48,813 (40.7) | 3,820 (40.8) | 1,967 (49.0) | <0.001 |
| Hyperlipidemia, n (%) | 86,113 (71.9) | 5,531 (59.1) | 2,557 (63.7) | <0.001 |
| Hypertension, n (%) | 99,121 (82.7) | 6,964 (74.4) | 3,157 (78.7) | <0.001 |
| Atrial fibrillation, n (%) | 7,107 (5.9) | 350 (3.7) | 153 (3.8) | <0.001 |
| Sleep apnea, n (%) | 24,923 (20.8) | 1,803 (19.3) | 988 (24.6) | <0.001 |
| COPD, n (%) | 26,541 (22.1) | 1,702 (18.2) | 859 (21.4) | <0.001 |
| Substance use, n (%) | 8,877 (7.4) | 941 (10.1) | 517 (12.9) | <0.001 |
| BP medications, n (%) | 105,256 (87.8) | 7,404 (79.1) | 3,235 (80.6) | <0.001 |
| Anticoagulation, n (%) | 5,859 (4.9) | 357 (3.8) | 176 (4.4) | <0.001 |
| Statin, n (%) | 69,104 (57.7) | 4,068 (43.5) | 1,769 (44.1) | <0.001 |
| Aspirin, n (%) | 18,701 (15.6) | 1,290 (13.8) | 601 (15.0) | <0.001 |
| Current smoker, n (%) | 35,010 (29.2) | 3,127 (33.4) | 1,633 (40.8) | <0.001 |
| ALT (mg/dL), mean (SD) | 36.9 (23.9) | 46.0 (36.2) | 48.0 (36.2) | <0.001 |
| AST (mg/dL), mean(SD) | 29.1 (26.8) | 33.4 (31.2) | 35.0 (28.8) | <0.001 |

**Abbreviations:** HDL, high-density lipoprotein cholesterol; TG, triglycerides; HbA1c, hemoglobin A1c; BMI, body mass index; CAD, coronary artery disease; CHF, congestive heart failure; PAD, peripheral arterial disease; COPD, chronic obstructive pulmonary disease; BP, blood pressure; Statin, HMG-CoA-Reductase inhibitor; ALT, alanine aminotransferase; AST, aspartate aminotransferase

**Table 3.**
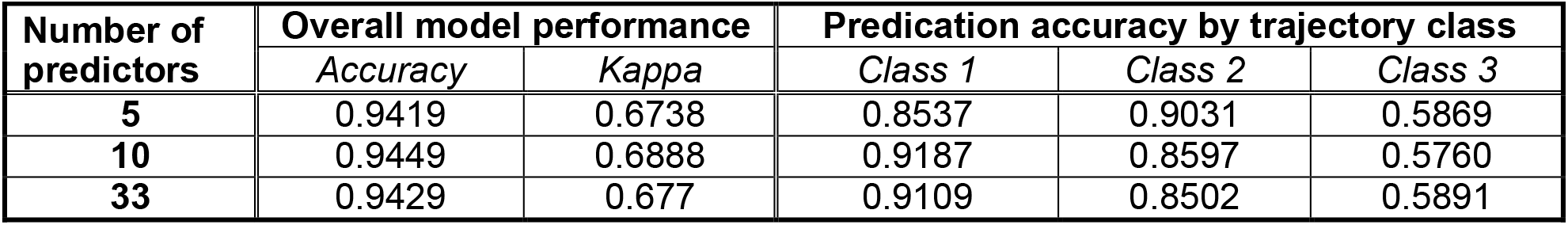
External validation of metformin glycemic response prediction model performance using random forests.

## Discussion

Currently, attainment of glycemic targets for individuals with type 2 diabetes is poor despite effective treatment options^2,3^. In this retrospective analysis of glycemic trajectories among individuals newly diagnosed with diabetes and treated with metformin monotherapy, we found that a substantial majority achieved glycemic control within 6 months of medication initiation. Fewer than 5% of individuals exhibited a glycemic non-response phenotype to metformin treatment. Furthermore, our results suggest that the process used by clinicians for selecting individuals for initial metformin monotherapy is effective, including a small proportion of individuals with a baseline HbA1c >11% at the time of treatment initiation. The results of our study highlight the glycemic effectiveness of metformin in the diabetes treatment armamentarium. Although clinical characteristics at baseline were not sufficient to predict the metformin non-response phenotype, glycemic response to metformin was evident within six months, suggesting that inadequate glucose control with metformin monotherapy can be identified quickly and easily with guideline-recommended glycemic surveillance. By enabling guideline-directed glycemic control in over 90% of individuals for whom initial metformin monotherapy was considered appropriate, metformin use may allow greater attention to blood pressure control and cholesterol lowering, the non-glycemic keys to diabetes management to mitigate cardiovascular disease risk^18^.

The results of this study employing real-world data extend evidence regarding initial diabetes treatment derived from the ADOPT and UKPDS clinical trials. The recent expansion in non-insulin diabetes medication options have shifted the focus to therapy for those with type 2 diabetes and excess cardiorenal risk or obesity. However, a great unmet need in global diabetes management is adequate early and durable glycemic control on metformin or other first-line diabetes treatments^19,20^. Unmet glycemic targets remain an urgent issue, often exacerbated by therapeutic inertia. Despite an explosion of glucose lowering medications over the last two decades, the achievement of glycemic control targets remains low. While the advent of treatments with cardiovascular, renal, and metabolic benefits is poised to revolutionize type 2 diabetes care, early glycemic control remains an effective, low-cost intervention that can reduce risk of complications. Indeed, UKPDS demonstrated that initial glucose control after diabetes diagnosis represents a critical period with long-term impact on clinical outcomes^5^. The results of this study suggest that with guideline-directed treatment and surveillance, many people with type 2 diabetes can achieve glycemic goals for multiple years with metformin alone.

With greater type 2 diabetes treatment options, many recent studies have focused on individuals requiring additional therapies after failure of first-line treatment. Such evidence supporting use of newer medications is essential but inadvertently ignores a large percentage of patients who have controlled glycemic levels on metformin. For example, the recently completed Glycemia Reduction Approaches in Type 2 Diabetes: A Comparative Effectiveness Study (GRADE) compared four glucose-lowering therapies as second-line treatment among adults with HbA1c of 6.8-8.5% on metformin monotherapy^21-23^. The GRADE study provided valuable comparative data on second-line treatments but does not inform initial treatment selection. Similarly, recent studies utilized real-world data to develop treatment selection models among second-line glucose-lowering treatments^24-27^. These studies provide critical evidence towards personalized treatment decision-making in diabetes care but do not address what may be the most important diabetes treatment choice: how to achieve effective early glycemic control. Our results suggest metformin alone is sufficient for A1c reduction in most individuals with newly diagnosed diabetes for whom single-drug treatment is thought to be appropriate.

This study of metformin glycemic response phenotypes falls at the intersection of two areas of mounting interest in diabetes research and care: type 2 diabetes heterogeneity and precision medicine. Following a landmark study that stratified individuals with newly diagnosed diabetes into 6 “clusters”^28^, many studies across a variety of clinical contexts have replicated statistically distinguishable subtypes of type 2 diabetes^29,30^. However, insights into type 2 diabetes subtypes based on clinical characteristics measured at the time of diagnosis have yet to translate to precise strategies for type 2 diabetes treatment^29,30^. Our study complements prior work to subclassify type 2 diabetes early after diagnosis by defining distinct metformin response phenotypes. However, clinical precision medicine tools to guide initial type 2 diabetes treatment choice remain elusive as we were not able to predict non-response to metformin with the data available in this study – a crucial next step towards tailoring initial glucose-lowering treatment in individuals newly diagnosed with diabetes.

Our study has important limitations. First, we limited the study sample to individuals treated with metformin monotherapy after diabetes diagnosis and censored individuals when they started a second or alternative diabetes treatment. This study design choice focused our analysis on the motivating question – to describe glycemic trajectories in response to metformin treatment – at the expense of more broadly describing early glucose control trajectories across the spectrum of treatment choices made early after diabetes diagnosis. Second, the accuracy of the prediction model for classifying individuals with high initial HbA1c who achieved early glycemic control may be inflated by the selection of individuals treated with metformin monotherapy in the study. In other words, this retrospective analysis included only individuals selected by providers to receive metformin monotherapy, a choice that may have been influenced by unmeasured factors suggesting a potentially brisk glycemic response among patients who would often initially be treated with multiple medications. Additionally, this study does not address glycemic efficacy in individuals for whom initial treatment choices include SGLT2i or GLP-1RA based on cardio-renal-metabolic risk. Finally, the study sample of Veterans was largely older men, which may limit generalizability to other populations. In subgroup analyses of the ADOPT study, however, the association of rosiglitazone compared to metformin with monotherapy failure was weaker in younger (≤50 years) rather than older individuals and demonstrated no heterogeneity by sex^8^.

In conclusion, we identified distinct glycemic response trajectories after initiation of metformin monotherapy as initial type 2 diabetes treatment. The vast majority of individuals treated with metformin alone achieved adequate glycemic control in the first two years of treatment, and this was sustained over five years in most. In light of the pervasive failure to meet glycemic targets, these data inspire implementation strategies for metformin initiation at diagnosis of type 2 diabetes. Critically, the trajectories were evident within six months of treatment initiation, suggesting that guideline recommended glycemic surveillance would allow early identification of individuals with an inadequate glycemic response to metformin. Routinely collected clinical data were not sufficient to accurately predict a metformin non-response phenotype, highlighting the challenge of translating type 2 diabetes sub-phenotypes into precision medicine tools. Taken together, our results support metformin’s placement as an effective initial type 2 diabetes treatment and highlight the role of routine care processes such as glycemic surveillance in achieving glycemic control in the critical early period after diabetes diagnosis.

## Supporting information

Supplemental Results

STROBE checklist

## Data Availability

Code for all statistical analysis is available upon request. A deidentified, anonymized limited data set derived from the datasets used for the analysis can be made available upon reasonable request from researchers with necessary human subjects research oversight and in accordance with VA data sharing policies.

## Funding statement

The funders had no role in the conduct of the study. This publication does not represent the views of the Department of Veteran Affairs or the United States Government. S.R. was supported by VA award IK2-CX001907, VA award I01-BX006417, a Webb-Waring Biomedical Research Award from the Boettcher Foundation, and US National Institutes of Health award 1P30DK116073. J.E.B.R. was supported by US National Institutes of Health award 1P30DK116073 and VA CX001532.

## Conflict of interest

The authors report no conflicts of interest relevant to this study.

## Author contributions

S.R., W.G.L., D.G., and J.E.B.R. designed the study. W.G.L. and T.W. conducted the analyses. S.R., W.G.L., and M.R.H. drafted the manuscript. All authors reviewed and provided critical revisions to the manuscript. The corresponding author attests that all listed authors meet authorship criteria and that no others meeting the criteria have been omitted. The lead author (S.R.) is the guarantor of the work and affirms that the manuscript is an honest, accurate, and transparent account of the study being reported with no important or intentional omissions.

## Copyright/license for publication

The Corresponding Author has the right to grant on behalf of all authors and does grant on behalf of all authors, a worldwide license to the Publishers and its licensees in perpetuity, in all forms, formats and media (whether known now or created in the future), to i) publish, reproduce, distribute, display and store the Contribution, ii) translate the Contribution into other languages, create adaptations, reprints, include within collections and create summaries, extracts and/or, abstracts of the Contribution, iii) create any other derivative work(s) based on the Contribution, iv) to exploit all subsidiary rights in the Contribution, v) the inclusion of electronic links from the Contribution to third party material where-ever it may be located; and, vi) license any third party to do any or all of the above.

## Notes

### Competing Interest Statement

The authors have declared no competing interest.

### Author Declarations

The Colorado Multiple Institutional Review Board and the Research & Development Committee of the VA Eastern Colorado Healthcare System provided human subjects oversight and approval for this work.

