## Supplemental Results for "Glycemic response trajectories on metformin monotherapy in real-world diabetes care"

**<u>Online-only Supplement</u>**

**Supplemental Table 1.** Predictor variables used in metformin response prediction model development.

**Supplemental Table 2.** Trajectory model fit with three to six trajectory classes.

**Supplemental Table 3.** Trajectory class size when fitting three to six classes.

**Supplemental Figure 1.** Glycemic response trajectories for four-, five-, and six-class models.

**Supplemental Table 4.** Reasons for censoring by trajectory class.

**Supplemental Figure 2.** Two-year glycemic trajectories with three classes when metformin adherence was not required for inclusion.

**Supplemental Table 5.** Variable importance for predicting trajectory class membership.

**Supplemental Table 6.** Overall prediction accuracy across different model development approaches.

**Supplemental Table 7.** Prediction accuracy by trajectory class across different model development approaches.

**Supplemental Table 1.** Predictor variables included in metformin response prediction model development.

| Model | Predictor |
|---|---|
| 5-variable | Age |
| | Hemoglobin A1c |
| | Body mass index |
| | High density lipoprotein cholesterol |
| | Triglycerides |
| 10-variable | Heart failure |
| | Coronary artery disease |
| | Liver disease |
| | Alanine aminotransferase |
| | Aspartate aminotransferase |
| 33-variable | Sex |
| | Cancer |
| | Kidney disease |
| | Amputation |
| | Stroke |
| | Peripheral artery disease |
| | Hepatitis |
| | Dialysis |
| | Anemia |
| | Arthritis |
| | Fatigue |
| | Depression/Anxiety |
| | Hyperlipidemia |
| | Hypertension |
| | Atrial fibrillation |
| | Sleep apnea |
| | Chronic obstructive pulmonary disease |
| | Substance use disorder |
| | Blood pressure medication use |
| | Anticoagulation medication use |
| | HMG-CoA Reductase inhibitor (statin) use |
| | Aspirin use |
| | Current smoker |

**Supplemental Table 2.** Trajectory model fit with three to six trajectory classes.

| | 3 Classes | 4 Classes | 5 Classes | 6 Classes |
|---|---|---|---|---|
| **Bayes Information Criteria (BIC)** | 1196482 | 1175746 | 1165076 | 1158391 |

Note: lower BIC implies better model fit to the observed data.

**Supplemental Table 3.** Trajectory class size as a proportion of total sample size (%) when fitting three to six classes.

| Model | Group 1 | Group 2 | Group 3 | Group 4 | Group 5 | Group 6 |
|---|---|---|---|---|---|---|
| 3 Classes | 7.14 | 89.72 | 3.14 | - | - | - |
| 4 Classes | 1.55 | 88.46 | 6.87 | 3.11 | - | - |
| 5 Classes | 2.23 | 5.79 | 87.98 | 2.66 | 1.35 | - |
| 6 Classes | 1.80 | 0.98 | 87.14 | 5.87 | 2.83 | 1.38 |

Note: Group numbers in this table do not correspond to the three Class labels described in the main manuscript for the three-class model.

**Supplemental Figure 1.** Glycemic response trajectories for four-, five-, and six-class models.

**A. Four-class model.**

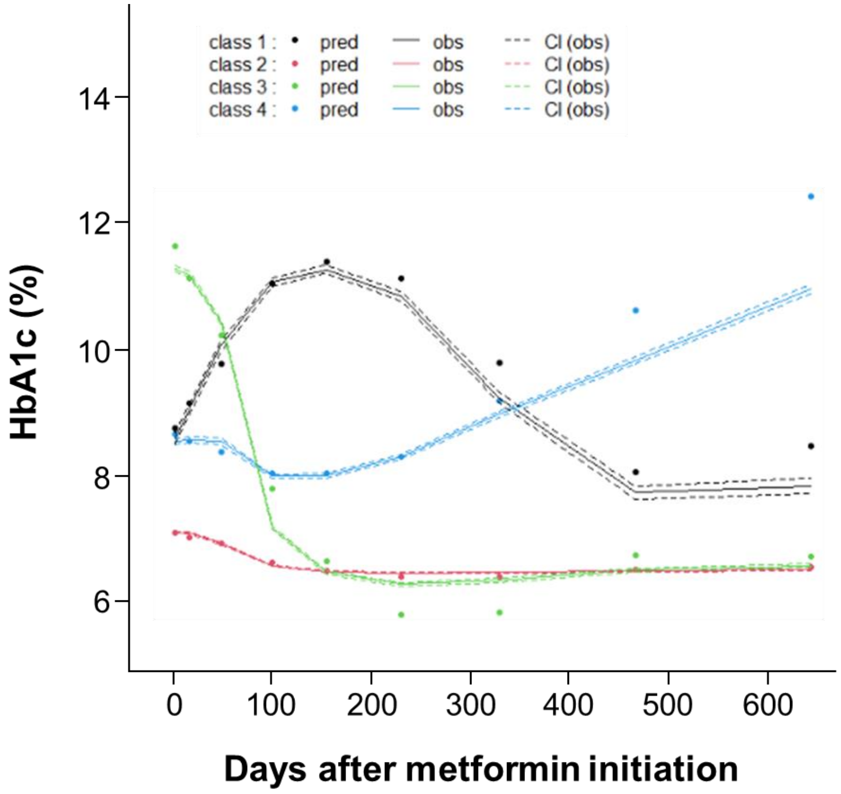


**B. Five-class model.**

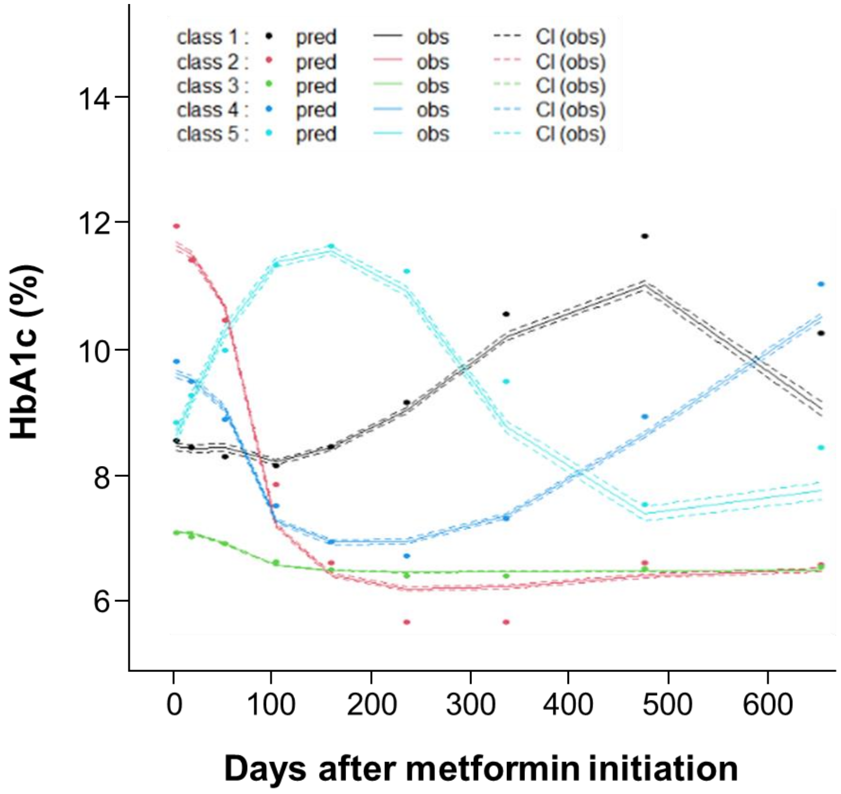

**C. Six-class model.**

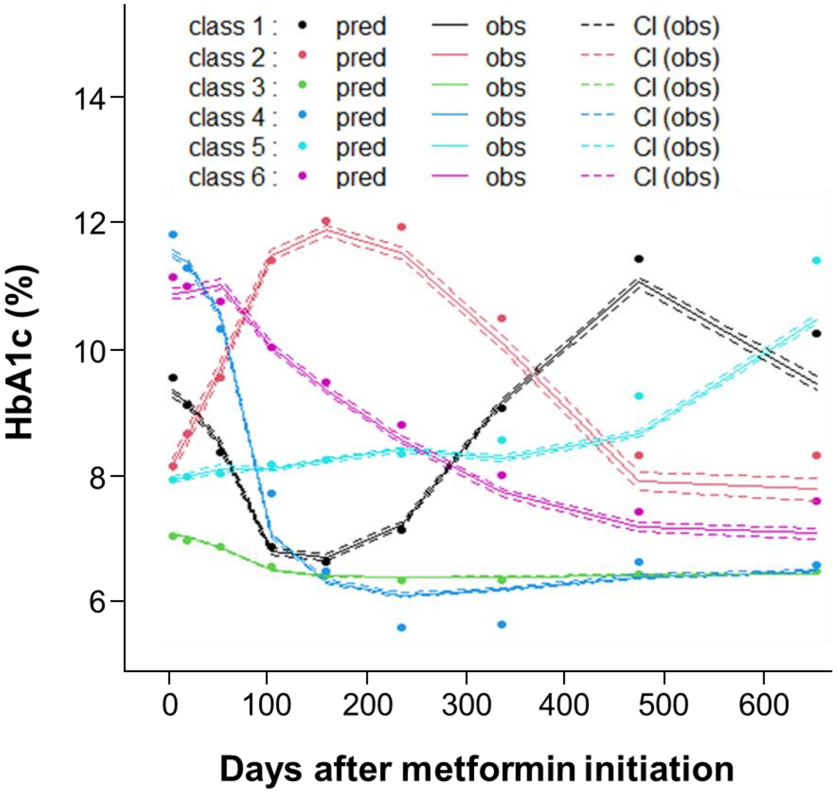

**Supplemental Table 4.** Reasons for censoring by trajectory class.

| Censoring reason | Trajectory Class | 6 months N (%) | 12 months N (%) | 18 months N (%) | 24 months N (%) |
|---|---|---|---|---|---|
| Any | Class 1 | 10,091 (8.0) | 58,882 (46.7) | 67,876 (53.9) | 79,600 (63.2) |
| | Class 2 | 1,455 (14.5) | 5,011 (50.0) | 5,823 (58.1) | 6,789 (67.8) |
| | Class 3 | 1,803 (40.9) | 3,618 (82.0) | 4,112 (93.2) | 4,334 (98.3) |
| Death | Class 1 | 177 (0.1) | 617 (0.5) | 1,029 (0.8) | 1,448 (1.2) |
| | Class 2 | 19 (0.2) | 48 (0.5) | 77 (0.8) | 93 (0.9) |
| | Class 3 | 5 (0.1) | 11 (0.3) | 12 (0.3) | 14 (0.3) |
| Initiation of 2nd diabetes medication | Class 1 | 4,481 (3.6) | 8,750 (7.0) | 11,740 (9.3) | 14,390 (11.4) |
| | Class 2 | 1,002 (10.0) | 1,431 (14.3) | 1,757 (17.5) | 2,031 (20.3) |
| | Class 3 | 1,594 (36.1) | 2,588 (58.7) | 3,017 (68.4) | 3,199 (72.5) |
| Metformin adherence <80% | Class 1 | 5,157 (4.1) | 48,790 (38.7) | 54,059 (42.9) | 62,461 (49.6) |
| | Class 2 | 413 (4.1) | 3,474 (34.7) | 3,907 (39.0) | 4,559 (45.5) |
| | Class 3 | 194 (4.4) | 1,000 (22.7) | 1,062 (24.1) | 1,100 (24.9) |
| Admission to long-term care residence | Class 1 | 276 (0.2) | 725 (0.6) | 1,048 (0.8) | 1,301 (1.0) |
| | Class 2 | 21 (0.2) | 58 (0.6) | 82 (0.8) | 106 (1.1) |
| | Class 3 | 10 (0.2) | 19 (0.4) | 21 (0.5) | 21 (0.5) |

**Supplemental Figure 2.** Two-year glycemic trajectories with three classes when metformin adherence was not required for inclusion.

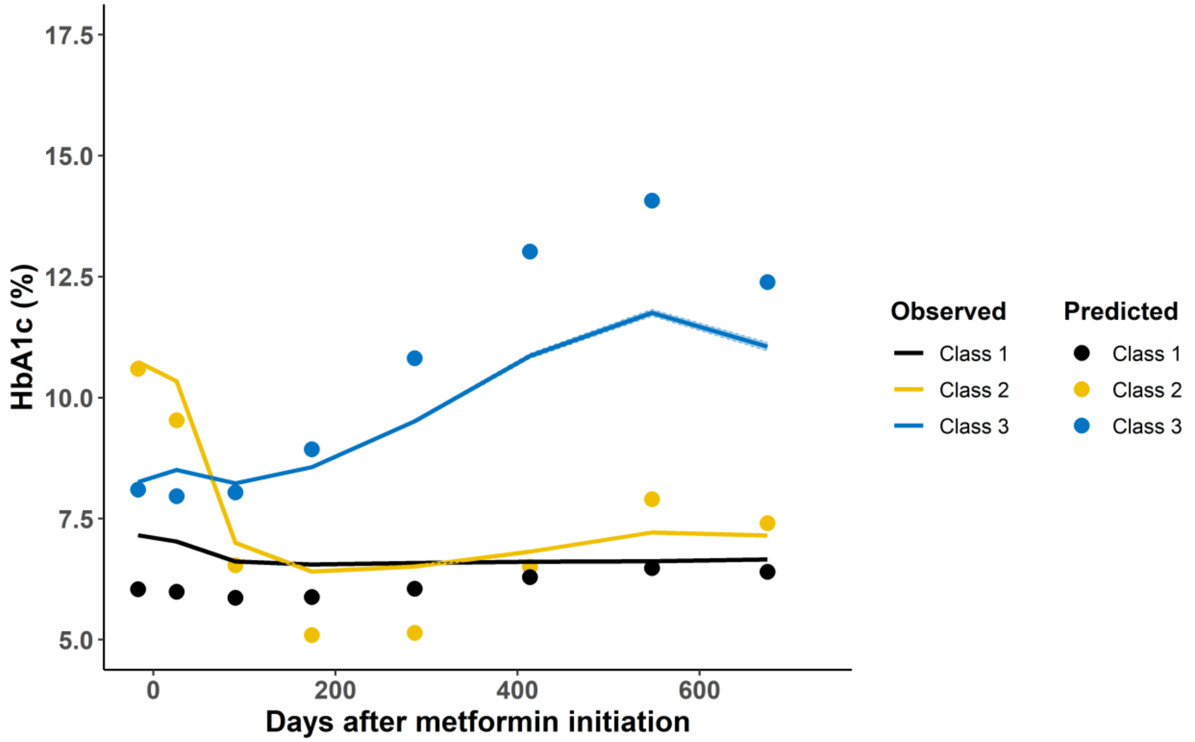

**Supplemental Table 5.** Variable importance for predicting trajectory class membership.

| Model | Variable | Random Forest | Neural Networks | Support Vector Machine |
|---|---|---|---|---|
| 5-variable | HbA1c | 100 | 100 | 100 |
| | Age | 7.915 | 21.968 | 17.166 |
| | BMI | 6.892 | 12.796 | 7.173 |
| | Triglycerides | 5.512 | 0 | 16.024 |
| | HDL cholesterol | 0 | 2.263 | 5.162 |
| 10-variable | HbA1c | 100 | 100 | 100 |
| | Age | 12.202 | 3.451 | 24.486 |
| | BMI | 11.249 | 17.896 | 15.183 |
| | Triglycerides | 11.402 | 0 | 24.622 |
| | HDL cholesterol | 8.315 | 1.885 | 13.734 |
| | Heart failure | 0.202 | 7.613 | 0.994 |
| | Coronary artery disease | 0.605 | 9.379 | 8.929 |
| | Liver disease | 0 | 69.72 | 0 |
| | Alanine aminotransferase | 0 | 5.77 | 16.279 |
| | Aspartate aminotransferase | 0 | 5.03 | 3.739 |
| 33-variable | HbA1c | 100 | NA[a] | NA[a] |
| | Age | 9.01 | | |
| | BMI | 8.367 | | |
| | Triglycerides | 8.152 | | |
| | HDL cholesterol | 5.815 | | |
| | Heart failure | 0 | | |
| | Coronary artery disease | 6.634 | | |
| | Liver disease | 0 | | |
| | Alanine aminotransferase | 6.075 | | |
| | Aspartate aminotransferase | 5.692 | | |
| | Sex | 0 | | |
| | Cancer | 0.61 | | |
| | Kidney disease | 0 | | |
| | Amputation | 0 | | |
| | Stroke | 0 | | |
| | Peripheral artery disease | 0 | | |
| | Hepatitis | 0 | | |
| | Dialysis | 0 | | |
| | Anemia | 0.359 | | |
| | Arthritis | 0.64 | | |
| | Fatigue | 0 | | |
| | Depression/Anxiety | 0.609 | | |
| | Hyperlipidemia | 0.478 | | |
| | Hypertension | 0.391 | | |
| | Atrial fibrillation | 0 | | |
| | Sleep apnea | 0.523 | | |
| | Chronic obstructive pulmonary disease | 0.464 | | |
| | Substance use disorder | 0 | | |
| | Blood pressure medication use | 0.442 | | |
| | Anticoagulation medication use | | | |
| | HMG-CoA Reductase inhibitor (statin) use | 0.523 | | |
| | Aspirin use | 0.472 | | |
| | Current smoker | 6.336 | | |

[a] 33-variable model was attempted only with random forests.

**Supplemental Table 6.** Overall prediction accuracy across different model development approaches.

| Method | 5-variable model | | 10-variable model | |
|---|---|---|---|---|
| | *Accuracy* | *Kappa* | *Accuracy* | *Kappa* |
| Random Forest | 0.9419 | 0.6738 | 0.9449 | 0.6888 |
| Neural Networks | 0.9435 | 0.6777 | 0.9434 | 0.6779 |
| Support Vector Machines | 0.9422 | 0.6588 | 0.943 | 0.6622 |

**Supplemental Table 7.** Prediction balanced accuracy by trajectory class across different model development approaches.

| Method | 5-variable model | | | 10-variable model | | |
|---|---|---|---|---|---|---|
| | *Class 1* | *Class 2* | *Class 3* | *Class 1* | *Class 2* | *Class 3* |
| Random Forest | 0.8537 | 0.9031 | 0.5869 | 0.9187 | 0.8597 | 0.5760 |
| Neural Networks | 0.8484 | 0.9291 | 0.5526 | 0.9253 | 0.8513 | 0.5542 |
| Support Vector Machines | 0.8245 | 0.9129 | 0.5451 | 0.9152 | 0.8274 | 0.5364 |
